# Downsizing food: A systematic review and meta-analysis examining the effect of reducing served food portion sizes on daily energy intake and body weight

**DOI:** 10.1101/2021.09.22.21263961

**Authors:** Eric Robinson, India McFarland-Lesser, Zina Patel, Andrew Jones

## Abstract

**Background:** Portion sizes of many foods have increased over time and reducing food portion sizes has been proposed as a public health strategy to reduce obesity. However, the extent to which reducing food portion sizes affects daily energy intake and body weight is unclear.

**Objective:** To systematically review and meta-analyse experimental studies that have examined the effect that serving smaller vs. larger portion sizes has on total daily energy intake.

**Design:** We used systematic review methodology to search identify eligible articles that used an experimental design to manipulate portion size served to human participants and measured energy intake for a minimum of one day. Multi-level meta-analysis was used to used to pool effects of portion size on daily energy intake.

**Results:** Fourteen eligible studies were included and 85 effects were included in the primary meta-analysis. There was a moderate-to-large reduction in daily energy intake when comparing smaller vs. larger portions (SMD = -.709 [95% CI: -.956 to -.461], p < .001, I2 = 80.6%) and evidence of a dose dependent response. Larger reductions to portion size and reducing portion sizes of multiple meals per day both resulted in larger decreases in daily energy intake. There was also evidence of a curvilinear relationship between portion size and daily energy intake, whereby reductions to daily energy intake were markedly smaller when reducing portion size from very large portions. In a subset of studies that measured body weight (n=5), being served smaller portions was associated with less weight gain than larger portions (SMD = .536 ([95% CI: .268 to .803], p < .001, I^2^ = 47.0%).

**Conclusions:** Smaller food portion sizes substantially decrease daily energy intake and there is evidence that over time this results in lower body weight. Reducing food portion sizes may be an effective population level strategy to reduce obesity.

## Introduction

Large portion sizes of commercially available food products have been identified as a likely contributor to the rise in overweight and obesity observed across most of the developed world (1-3). In particular, there is evidence that food portion sizes have increased over time, with the current food environment characterised by a wide availability of energy dense food products sold in portion sizes that promote excessive energy intake (2, 4-6). There is also now a consistent body of evidence indicating that manipulating the portion size of a meal served affects acute energy intake during that meal in children and adults (7-9). A recent meta-analysis of short-term studies estimated that doubling the served portion size at a meal increases meal energy intake by 35%, on average (10). Based on these findings, public health measures to reduce portion sizes of food and drink products have been proposed as a potentially effective intervention to reduce obesity (11).

Yet, the long-term effects of reducing food portion sizes are less clear. Recent findings indicate that smaller portion sizes may ‘normalize’ overtime and be accepted by consumers (12-14). However, less research has examined whether consumers ‘compensate’ for reduced portion sizes by eating more at later meals, or whether reductions in portion size instead lead to a meaningful decrease in daily energy intake and in doing result in longer-term reductions to body weight (5, 15). One study that examined compensatory eating found decreasing the portion size of a main course served at lunch resulted in decreased energy intake from the main course, but resulted in an increase in amount of energy consumed at dessert (16). Lewis et al., examined the effect of reducing breakfast portion size relative to a larger portion size but found no difference in total daily energy intake between portion size conditions (17). Conversely, other studies have found that serving smaller relative to larger served portion sizes resulted in lower daily energy intake over multiple days (18, 19). If smaller portion sizes do decrease daily energy intake it is also unclear what the approximate size of this relationship is likely to be (i.e. reducing portion size by 100kcals results in *x*kcals reduction in daily energy intake). Research examining the impact that portion size manipulations have on body weight have produced mixed findings to date, which may be due to studies having relatively short follow-up durations or lacking sufficient statistical power to detect relatively modest changes in body weight attributable to portion size manipulations (20-22). Therefore, the aims of the present research were to systematically review and meta-analyse the impact that experimentally manipulating portion size has on total daily energy intake and changes in body weight.

## Method

### Eligibility criteria and study selection

We included studies that used an experimental design to directly manipulate the portion size of food served to participants and measured energy intake across the course of at least one day.

#### Participants

Studies of human participants were eligible. Studies that sampled participants with a diagnosed medical/chronic health condition or currently undergoing treatment which may influence appetite (e.g. diabetes, bariatric surgery patients) were not eligible. There were no other exclusion criteria based on participant characteristics.

#### Intervention

Studies were required to have manipulated portion sizes (i.e. amount of food served to participants, also known as ‘serving size’) provided to participants. Studies that manipulated portion size of a single food/meal were eligible, as were studies that manipulated all foods/meals served across the day. Studies that only reduced portion size of drink(s) were not eligible, as our focus was on food. However, if a study manipulated food and drinks it was deemed eligible. Studies were required to serve or provide all participants with the same food type and to have achieved different portion size conditions by only altering the weight/volume of food served.

#### Intervention (smaller portion sizes) vs. comparator (larger portion sizes) conditions

In studies with two portion size conditions the ‘comparator’ condition was the larger portion size condition and the ‘intervention’ condition was the smaller portion size condition. Some studies described their manipulation as examining effect of ‘larger’ portion sizes vs. ‘standard’ portion sizes on energy intake, and so to ensure consistency with the above conceptualisation, we treated the larger condition as the comparator condition. Studies with multiple portion size conditions (e.g. 100% vs. 75% vs. 50%) were eligible and contributed multiple effects to the present review (e.g. 100% vs. 75%, 75% vs. 50%, 100% vs. 50%).

#### Outcomes

Eligible studies were required to have measured energy intake across the course of a minimum of one day. Studies that measured energy intake through objective researcher measurement (e.g. weighing of food pre/post eating), participant self-reported (e.g. dietary recall data), or a combination of the two were eligible.

#### Study Design

Studies that adopted a within-subjects/repeated measures design (i.e. participants receive both smaller and larger portions) were eligible, as were between-subjects designs (i.e. participants were randomized to receive either the smaller vs. larger portions). Studies that measured energy intake in controlled laboratory and in real-world settings were eligible. Studies that required participants to consume a meal or food in full (e.g. compulsory consumption of a set amount of breakfast) were eligible. Studies that ‘crossed’ a portion size manipulation with another study manipulation (e.g. manipulation of both portion size and energy density of food served in the same study) were eligible, although only contrasts between portion size conditions were included in analyses. For studies that did not manipulate all meals/foods (e.g. only manipulating portion size of lunch), eligible studies were required to measure and report energy intake at that meal(s) that energy portion size was manipulated, in order to quantify the effect of the portion size manipulation independent of non-manipulated foods/meals.

### Article identification strategy

In September 2020, we searched PsycINFO, PubMed and SCOPUS (from date of inception onwards) using combinations of search terms relating to portion size and energy intake (see online supplementary materials for an example). To identify further published literature, we used a snowballing approach by searching the reference lists of eligible papers and by contacting authors to ask whether they had authored any other potentially eligible studies. To identify grey literature, we conducted additional searches of the OSF preprint archive (a database covers 30 other preprint archives, including PsychArxiv and Nutrixiv). Two authors independently screened and judged eligibility of articles identified through electronic searches. A single author identified potentially eligible articles using the snowballing and grey literature approaches, and all potentially eligible articles were verified by a second independent author. Discrepancies for eligibility were resolved by discussion or were adjudicated by a third author.

### Data extraction

Two authors extracted the following information and any extraction discrepancies were resolved through discussion or a third author adjudicated; study sample information (e.g. country of study, participant group sampled, summary information on participant demographic characteristics, exclusion criteria for participant eligibility), portion size manipulation information (e.g. foods/meals manipulated, number of kcals served in portion size conditions, total number of kcals served per day in portion size conditions), study design information (e.g. within-subjects vs. between-subjects design), measurement of energy intake (self-reported vs. researcher measured), use ad-libitum intake vs. compulsory intake (i.e. whether any meals were required to be eaten in full as part of the method), number of days energy intake was measured for, energy intake information (e.g. energy intake from portion size manipulated meals, non-manipulated meals and total daily energy intake, and correlation between comparator vs. intervention energy intake), results of any participant characteristic moderation analyses reported (e.g. does effect of portion size on energy intake differ in normal weight vs. participant with obesity?), whether body weight was measured before and after each comparator vs. intervention condition, and risk of bias indices (see below).

### Risk of bias indicators

Informed by guidelines for best practice when conducting randomized control trials and experimental studies of eating behaviour (23-26), studies were coded for nine risk of bias indicators. Studies that relied on self-reported energy intake (as opposed to researcher measured), did not use key participant exclusion eligibility criteria (e.g. use of medication affecting appetite, currently pregnant), were missing key methodological details (e.g. no information provided about foods used), did not report use of random allocation to conditions, required participants to consume some meals/food in full (e.g. compulsory consumption of smaller vs. larger portion sizes), did not address demand characteristics (e.g. no attempt to blind participants to study aims or check if participants were aware of study aims), had a small sample size (N<12 for within-subject studies and N < 20 per condition for between-subjects studies), did not pre-register the study or failed to report information on conflicts of interest statement (or reported a relevant conflict) were considered higher in risk of bias.

### Analyses

Pre-registered analyses and study data are available online: https://osf.io/dj4yf/. Authors were contacted and asked to provide details if statistical information required for analyses examining energy intake or body weight outcomes was missing. No within-subject/repeated measures studies reported the correlation between daily energy intake in the larger vs. smaller portion size conditions. As this information was required to calculate measures of effect size in within subject designs, we contacted all study authors to request this information and calculated the average (r = 0.8). As only a minority of authors provided this information, in sensitivity analyses we examined if results of meta-analyses differed based on the magnitude of the correlation (including r = 0.4 and r = 0.6). Any deviations from planned analyses are reported in the online supplemental material.

#### Primary analyses

##### Effect of portion size condition on daily energy intake

In a primary model we examined the effect of portion size condition (smaller vs. larger) on daily energy intake for all included studies. Because individual studies contributed multiple portion size comparisons, we used multi-level meta-analysis to account for the dependency of these effects (27). We defined outliers as any effect sizes for which the upper bound of the 95% confidence interval was lower than the lower bound of the pooled effect confidence interval (i.e., extremely small effects) or for which the lower bound of the 95% confidence interval was higher than the upper bound of the pooled effect confidence interval (extremely large effects), using standardised effects. We identified influential cases as any effects with DFBETA values > 1 (indicative of a >1 change in the standard deviation of the estimated co-efficient after removal) (28). We conducted Egger’s test (29) and a trim and fill procedure (30) to examine potential publication bias. See online supplementary materials for detailed information on outlier, influential case and publication bias analyses. If we identified any outliers they were removed in all subsequent analyses on daily energy intake. For all meta-analyses we calculated the standardised mean difference as a measure of effect size and SMDs of 0.2, 0.5 and 0.8 are typically considered small, moderate, and large sized effects (31). To aid interpretation, where appropriate we also meta-analysed and present mean difference in energy intake (kcals) between portion size conditions.

##### Participant and study features: effects on daily energy intake

We conducted sub-group analyses to examine if results differed between effects drawn from female vs. male samples and between studies that manipulated portion size at a majority of meals during the day (>2 meals) vs. fewer meals (≤ 2 meals). We planned to examine other participant characteristics (e.g. normal weight vs. overweight) in sub-group analyses but were unable to because too few studies reported sufficient data. Studies were variable in the number of days that they measured energy intake, and some studies reported effects on daily energy intake for each day of the study duration (effect of portion size on energy intake for day 1, 2, 3 etc.), so meta-regression was used to examine whether the impact of portion size on daily energy intake differed based on number of days energy intake was assessed for. One study examined energy intake at a 6-month follow-up; as this was a much longer follow-up period compared to the other studies, we excluded this data point from the meta-regression (although results were consistent with its inclusion). We also used meta-regression to examine if the effect of portion size on daily energy intake was related to the % magnitude of portion size reduction (i.e. smaller portion being 50% reduced compared to the larger portion) and difference in kcal served between the two portion size conditions.

##### Risk of bias indicators: effects on daily energy intake

We conducted sub-group analyses to examine if results from the primary analyses differed based on measurement of energy intake (researcher measured only vs. use of self-report), whether studies required participants to consume any meals in full (yes vs. no), use of random allocation to portion size conditions (yes vs. no) and whether demand characteristics were addressed in the study (yes vs. no).

#### Secondary analyses

##### Compensation effects

A sub-set of studies did not manipulate portion size at every meal and reported energy intake during the manipulated and/or energy intake post-manipulated meal. In a series of analyses limited to these studies we meta-analysed the effect of portion size on daily energy intake, manipulated meal energy intake and post-manipulated meal energy intake, to quantify the extent to which acute changes in energy intake caused by reducing portion size (i.e. difference in energy intake between portion size conditions at manipulated meals) were later compensated for. In 3 studies, the manipulated meal was ‘fixed’ (i.e. eaten in full by all participants) resulting in a standard deviation of 0. In sensitivity analyses we imputed the SDs for these fixed meal as the average SD (as a proportion of the mean) calculated from the non-fixed meals (∼29%).

##### Curvilinear relationship

Previous research has suggested that there may be a curvilinear relationship between increases in portion size and energy intake (10, 32), whereby the effect that manipulating portion size has on energy intake is reduced at larger more extreme portion sizes (e.g. medium vs. large) compared to smaller portion sizes (e.g. small vs. medium). A sub-set of studies (n=5) included three portion size conditions (e.g. large, medium, small) with similar sized increments in served portion size (e.g. 250 vs. 500 vs. 750kcals). We meta-analysed these studies and examined whether the reduction from the largest portion size (e.g. large vs. medium contrast) produced a similar effect on daily energy intake as the same sized from reduction from the intermediate portion size (e.g. medium vs. small) using sub-group analysis.

##### Effect of portion size condition on body weight

For studies that also measured body weight change, we conducted generic variance inverse meta-analysis with change in body weight (difference in change in body weight between the large and small portion size condition) as the outcome variable. If studies had more than two portion size conditions, because relatively subtle changes in energy intake would be unlikely to have a detectable effect on body weight over the short duration of studies included, to maximise statistical power a-priori we included the smallest and largest portion size condition from each study in the meta-analysis.

## Results

### Study characteristics

A total of 14 studies were included in the review, see Figure 1 for study selection flowchart. Nine studies were from the US, 4 were from the UK, and 1 study was from Singapore. The majority of studies sampled from university staff/students and the local community (12/14). Nine studies sampled males and females, 4 sampled females only and 1 sampled males only. Twelve studies were in adults and 2 were in children. Of the 13 studies that reported mean BMI, for 9 studies mean BMI was within the normal BMI range (18.5-24.9) and 4 studies had a mean BMI above this range (BMI≥25). Thirteen studies used within-subjects designs (portion size manipulated within participants) and one used a between-subjects design (portion size manipulated between participants). The total number of participants in each study ranged from N=19 to N=172. Portion size was manipulated and energy intake measured for 1 day in 6 studies, between 2-11 days in 6 studies, in one study for 4 weeks and in another study for 6 months. Six studies manipulated portion size at a single meal and the remaining 8 studies manipulated portion size at multiple meals. See Table 1 for individual study information.

**Table 1.** Summary information on included studies

| Study | Country and sample | Sample characteristics | Study design information | Number of participants in analysis | Foods served | Portion size manipulation | Body weight measurement pre-post comparator and intervention conditions |
| --- | --- | --- | --- | --- | --- | --- | --- |
| Blatt 2012 (42) | USA<br>Local community & university | Men (N=28)<br>Age:<br>M = 26.8 years<br>BMI:<br>M = 24.9<br>Women (N=40)<br>Age:<br>M = 27.6 years<br>BMI:<br>M = 23.3 | Within-subjects<br>Researcher measured EI<br>EI measured for 1 day | N = 68<br>28M, 40F | PS of main entrée of all meals (breakfast, lunch dinner) manipulated<br>Sides and beverages not manipulated and consumed ad libitum<br>Compulsory eating of manipulated entrees | <u>Men</u><br><u>Smaller PS condition</u><br>1000 kcals served per day for manipulated meals<br>5337 kcals served per day<br>kcals eaten per day: M =2455, SE = 97<br><u>Larger PS condition</u><br>1570 kcals served per day for manipulated meals<br>5904 kcals served per day<br>kcals eaten per day: M = 2751, SE = 92<br><u>Women</u><br><u>Smaller PS condition</u><br>700 kcals served per day for manipulated meals<br>4146 kcals served per day<br>kcals eaten per day: M =1805, SE = 57<br><u>Larger PS condition</u><br>1100 kcals served per day for manipulated meals<br>4549 kcals served per day<br>kcals eaten per day: M = 1988, SE = 58 | Not measured |
| Fisher, 2007 (43) | USA | Children<br>Age: | Within-subjects | N = 116 | PS of main entrée of all meals | <u>Children</u><br><u>Smaller PS condition</u> | Not measured |
|  | Local community (Low-income family children) | M = 5 years<br>BMI:<br>M = 60 percentile<br>Gender:<br>24M, 35F<br>Mothers (N=58)<br>Age:<br>M = 30 years<br>BMI:<br>M = 34 | Researcher measured EI<br>EI measured for 1 day |  | (breakfast, lunch dinner) manipulated and afternoon snack<br>Sides and beverages not manipulated<br>No compulsory eating, all foods consumed ad libitum | 2727 kcals served per day<br>kcals eaten per day: M =1500, SD = 359<br><u>Larger PS condition</u><br>4006 kcals served per day<br>kcals eaten per day: M = 1639, SD = 378<br><u>Mothers</u><br><u>Smaller PS condition</u><br>4109 kcals served per day<br>kcals eaten per day: M =2819, SD = 502<br><u>Larger PS condition</u><br>5974 kcals served per day<br>kcals eaten per day: M = 2965, SD = 616 |  |
| French, 2014 (20) | USA<br>Large metropolitan medical complex employees | Age:<br>M = 42.6 years<br>BMI:<br>M = 29.8<br>Gender:<br>60M, 112F | Between-subjects<br>Self-report measured EI<br>EI measured 3 times per week at months 1,3 and 6 | N = 172 | PS of lunch manipulated<br>All other meals and snacks not manipulated (free-living)<br>No compulsory eating | <u>Small Lunch Box (~400kcal):</u><br>413 kcals served per manipulated meal (lunch)<br>kcals eaten per day: M = 1718, SE = 70<br><u>Medium Lunch Box (~800kcal):</u><br>821 kcals served per manipulated meal (lunch)<br>kcals eaten per day: M = 1792, SE = 68<br><u>Large Lunch Box (~1600kcal):</u><br>1604 kcals served per manipulated meal (lunch)<br>kcals eaten per day: M = 1996, SE = 71 | <u>Small lunch box</u><br>M kg = -0.1, SE = 0.43<br><u>Medium lunch box</u><br>M kg = -0.1, SE = 0.42<br><u>Larger lunch box</u><br>M kg = 1.1, SE = 0.44 |
| Gray, 2002 (44) | UK<br>Students and staff at a University | Age:<br>M = 24.3 years<br>BMI:<br>M = 22.6<br>Gender: 20M | Within-subjects<br>Researcher and self-report measured EI | N = 20 | PS of soup preload prior to lunch manipulated<br>Ad libitum consumption of pasta test meal | <u>Smaller PS condition</u><br>100 kcals served per manipulated meal (soup)<br>kcals eaten per day: M =2756.00, SE = 158.9<br><u>Larger PS condition</u><br>301 kcals served per manipulated meal (soup)<br>kcals eaten per day: M = 2798.40, SE = 155 | Not measured |
|  |  |  | EI measured for 1 day |  | (lunch). All other meals and snacks not manipulated (free-living)<br>Compulsory eating of soup preload |  |  |
| Haynes, 2020 (18) | UK<br>Local community and university | Age:<br>M = 31.6 years<br>BMI:<br>M = 26.0<br>37 % normal weight<br>63 % overweight/obesity<br>Gender:<br>15M, 15F | Within-subjects<br>Researcher measured EI<br>EI measured for 5 days | N = 30 | PS of lunch and dinner entrée manipulated<br>Ad-libitum eating of all other meals/sides/snacks<br>No compulsory eating | <u>Smaller than normal PS condition:</u><br>678 kcals served per day for manipulated meals (lunch and dinner entrees)<br>5074 kcals served per day<br>kcals eaten per day: M = 2238, SD = 490<br><u>Small-normal PS condition:</u><br>1086 kcals served per day for manipulated meals (lunch and dinner entrees)<br>5485 kcals served per day<br>kcals eaten per day: M = 2448, SD = 584<br><u>Large-normal PS condition:</u><br>1494 kcals served per day for manipulated meals (lunch and dinner entrees)<br>5897 kcals served per day<br>kcals eaten per day: M = 2543, SD = 592 | <u>Smaller than normal PS condition</u><br>M kg = -0.03, SD = 1.55<br><u>Small-normal PS condition</u><br>M kg = -0.03, SD = 1.34<br><u>Large-normal PS condition</u><br>M kg = 0.33, SD = 0.74 |
| Jeffery, 2007 (22) | USA<br>Local community and university | Age:<br>M = 33.0 years<br>BMI:<br>M = 28.9<br>Gender:<br>19F | Within-subjects<br>Self-report measured EI<br>EI measured 2 times per week for 4 | N = 19 | PS of lunch manipulated<br>Ad-libitum eating of lunch. All other meals and snacks not manipulated (free-living) | <u>Smaller PS condition</u><br>767 kcals served per meal<br>kcals eaten per day: M = 1875, SD = missing<br><u>Larger PS condition</u><br>1528 kcals served per meal<br>kcals eaten per day: M = 2153, SD = missing | <u>Smaller PS condition</u><br>M kg = 0.06, SD = 1.03<br><u>Larger PS condition</u><br>M kg = 0.64, SD = 1.16 |

|  |  |  | weeks |  | No compulsory eating |  |  |
| --- | --- | --- | --- | --- | --- | --- | --- |
| Kelly, 2009 (21) | UK<br>Local community and university | Men (N=21)<br>Age:<br>M = 29.7 years<br>BMI:<br>M = 25.3<br>43% normal weight<br>57% overweight/obesity<br>Women (N=22)<br>Age:<br>M = 31.7 years<br>BMI:<br>M = 23.7<br>68% normal weight<br>32% overweight/obesity | Within-subjects<br>Researcher measured EI<br>EI measured for 4 days | N = 43 | PS of all meals, snacks and drinks manipulated<br>Ad-libitum eating of all food and drink<br>No compulsory eating | <u>Men</u><br><u>Smaller PS condition</u><br>kcal served per day/meal - not reported<br>kcal eaten per day: M = 3490, SE = 220<br><u>Larger PS condition</u><br>kcal served per day/meal - not reported<br>kcal eaten per day: M = 4056, SE = 239<br><u>Women</u><br><u>Smaller PS condition</u><br>kcal served per day/meal - not reported<br>kcal eaten per day: M = 2721, SE = 137<br><u>Larger PS condition</u><br>kcal served per day/meal - not reported<br>kcal eaten per day: M = 2995, SE = 137 | MEN:<br><u>Smaller PS condition</u><br>M kg = 0.1, SD = missing (imputed as 1.2)<br><u>Larger PS condition</u><br>M kg = 0.9, SD = missing (imputed as 1.2)<br>WOMEN:<br><u>Smaller PS condition</u><br>M kg = 0.2, SD = missing (imputed as 1.2)<br><u>Larger PS condition</u><br>M kg = 0.6, SD = missing (imputed as 1.2) |
| Kral, 2004 (45) | USA<br>Local community and university | Age:<br>M = 23.4 years<br>BMI:<br>M = 23.1<br>Gender:<br>39F | Within-subjects<br>Researcher measured EI<br>EI measured for 1 day | N = 39 | PS of entrée at lunch manipulated<br>All foods and beverages were consumed ad libitum except for compulsory food | <u>500g condition:</u><br>750 kcal served per manipulated meal (lunch entrée)<br>kcal eaten per day: M = 1745, SE = 57<br><u>700g condition:</u><br>1050 kcal served per manipulated meal (lunch entrée) | Not measured |
|  |  |  |  |  | items with lunch<br>Compulsory eating of side dishes at lunch | kcal eaten per day: M = 1782, SE=51<br><u>900g condition:</u><br>1350 kcal served per manipulated meal (lunch entrée)<br>kcal eaten per day: M = 1855, SE = 54 |  |
| Lewis, 2015 (17) | UK<br>Local community & university | Age:<br>M = 42.5 years<br>BMI:<br>M = 29.0<br>Adults with overweight and obesity only<br>Gender:<br>15M/18F | Within-subjects<br>Researcher and participant measured EI<br>EI measured for 1 day | N = 33 | PS of breakfast meal manipulated<br>Ad-libitum eating of lunch and afternoon snack.<br>All other meals and snacks not manipulated (free-living)<br>Compulsory eating of breakfast | <u>40% reduction condition:</u><br>420 kcal served per manipulated meal (breakfast)<br>kcal eaten per day: M = 2190, SE = 104<br><u>20% reduction condition:</u><br>559 kcal served per manipulated meal (breakfast)<br>kcal eaten per day: M = 2365, SEM= 117<br><u>'Control' no reduction condition:</u><br>699 kcal served per manipulated meal (breakfast)<br>kcal eaten per day: M = 2459, SE = 94 | Not measured |
| McCrickerd, 2017 (46) | Singapore<br>Local community & university | Age:<br>M = 25.4 years<br>BMI:<br>M = 21.1<br>Gender:<br>53F | Within-subjects<br>Researcher and self-report measured EI<br>EI measured for 1 day | N = 51 | PS of breakfast meal manipulated<br>Ad-libitum eating of all meals<br>All other meals and snacks not manipulated (free-living)<br>No compulsory eating of meals | <u>Smaller PS condition</u><br>399 kcal served per manipulated meal (breakfast)<br>kcal eaten per day: M = 1668 kcal, SD = 521<br><u>Larger PS condition</u><br>599 kcal served per manipulated meal (breakfast)<br>kcal eaten per day: M = 1689kcal, SD = 489 | Not measured |
| Rolls 2006a 'Larger' (47) | USA | Men (N=16)<br>Age: | Within-subjects | N = 32 (16M, 16F) | PS of for all meals and snacks | <u>Men</u><br>kcal served per day - not reported / calculatable | Not measured |
|  | Local community & university | M = 24.4 years<br>BMI:<br>M = 24.7<br>Women (N=16)<br>Age:<br>M = 21.2 years<br>BMI:<br>M = 22.2 | Researcher measured EI<br>EI measured for 2 days |  | manipulated<br>Ad-libitum eating of all meals and beverages<br>Beverages were not manipulated<br>No compulsory eating of meals | kcal eaten per day:<br><u>100% condition</u>: M = 2964 kcal, SE = 133<br><u>150% condition</u>: M = 3461kcal, SE = 137<br><u>200% condition</u>: M =3774kcal, SE = 198<br><u>Women</u><br>kcal served per day - not reported / calculatable<br>kcal eaten per day:<br><u>100% condition</u>: M = 2188kcal, SE = 99<br><u>150% condition</u>: M = 2523kcal, SE = 99<br><u>200% condition</u>: M = 2717kcal, SE = 131 |  |
| Rolls 2006b 'Reductions' (48) | USA<br>Local community & university | Age:<br>M = 21.9 years<br>BMI:<br>M = 22.6<br>Gender:<br>24F | Within-subjects<br>Researcher measured EI<br>EI measured for 2 days | N = 24 | PS of all food manipulated<br>Ad-libitum eating of all meals and beverages<br>Beverages were not manipulated<br>No compulsory eating of meals | <u>Smaller PS condition</u><br>2846kcal served per day<br>kcal eaten per day: M =1951, SE = 65<br><u>Larger PS condition</u><br>3794kcal served per day<br>kcal eaten per day: M = 2207, SE = 67 | Not measured |
| Rolls, 2007 (19) | USA<br>Local community and university | Men (N=13):<br>Age:<br>M = 24.7 years<br>BMI:<br>M = 24.6<br>Women (N=10)<br>Age:<br>M = 25.8 years | Within-subjects<br>Researcher measured EI<br>EI measured for 11 days | N = 23 | PS of all meals (breakfast, lunch, dinner) and snacks manipulated<br>Ad libitum consumption of all meals (breakfast, lunch, dinner) and snacks) | MEN:<br><u>Smaller PS condition</u><br>4100 kcal served per day<br>kcal eaten per day: M = 2909, SE = 106<br><u>Larger PS condition</u><br>6150 kcal served per day<br>kcal eaten per day: M = 3328, SE = 114<br>WOMEN: | Not measured |
|  |  | BMI:<br>M = 22.9 |  |  | No compulsory eating | <u>Smaller PS condition</u><br>3400 kcals served per day<br>kcals eaten per day: M = 2073, SE = 97<br><u>Larger PS condition</u><br>5100 kcals served per day<br>kcals eaten per day: M = 2530, SE = 79 |  |
| Smethers, 2019 (49) | USA<br>Children from childcare centers | Age:<br>M = 4.4 years<br>BMI:<br>BMI-for-age percentile = 52.8<br>89% normal weight<br>11% overweight/obesity<br>Gender:<br>30M, 16F | Within-subjects<br>Researcher measured EI<br>EI measured for 5 days | N = 46 | PS of all food and beverages manipulated<br>Ad-libitum eating of all meals and beverages<br>No compulsory eating of meals | <u>Smaller PS condition</u><br>1627 kcals served per day<br>kcals eaten per day: M = 914, SE = 44<br><u>Larger PS condition</u><br>2450 kcals served per day<br>kcals eaten per day: M = 1081, SE = 44 | Not measured |

BMI = Body mass index, EI = energy intake, SE = standard error

**Figure 1.**
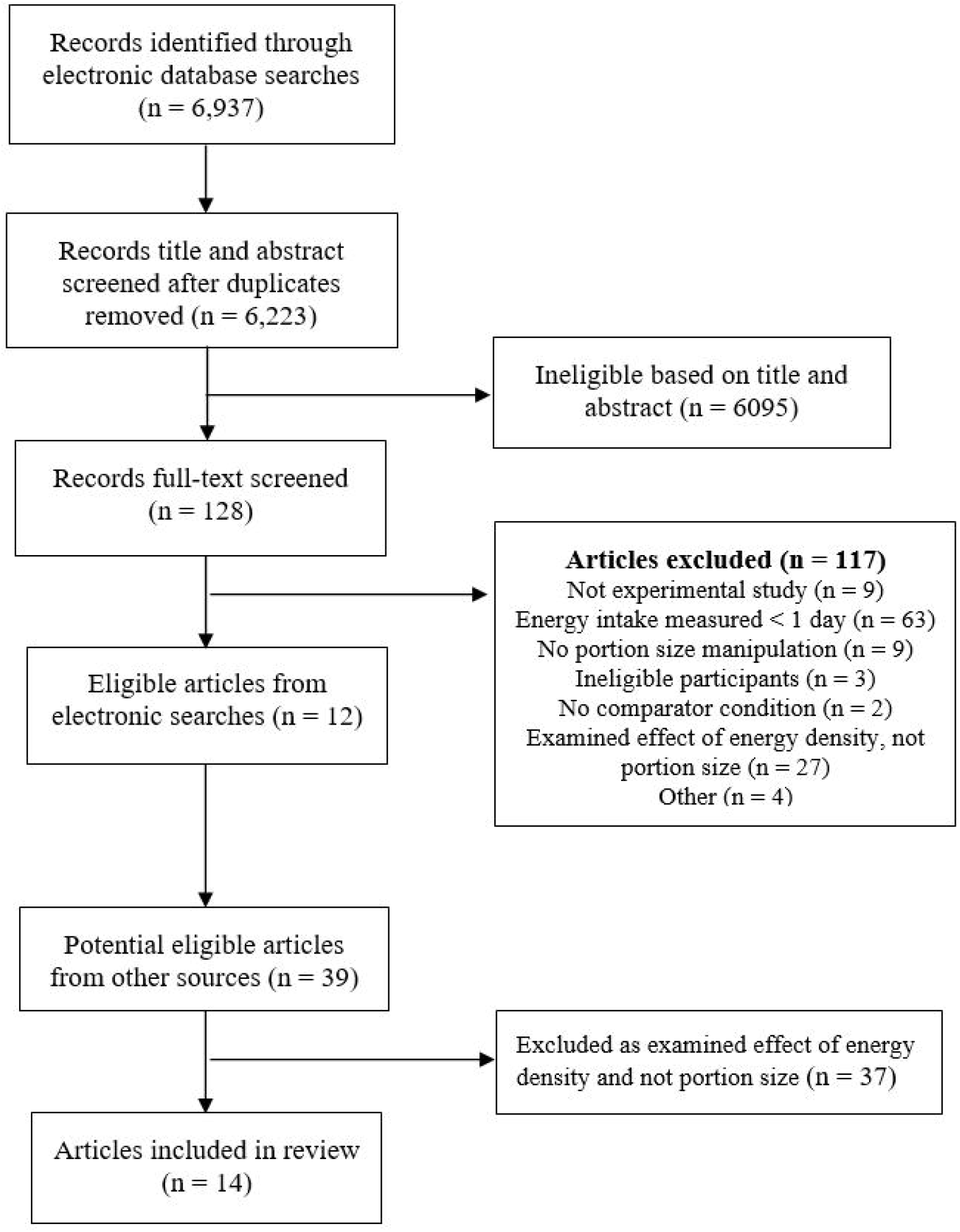
Study selection flowchart.

From the 14 studies, there were a total of 85 smaller vs. larger portion size daily energy intake comparisons, 35 of which were from female only samples, 23 male only and 27 were mixed sex. The size of portion size reduction examined (energy served in a larger portion condition vs. smaller portion condition) ranged from 20% to 74%, with a median of 33%. For the 65 portion size comparisons that the difference in kcals served between larger and smaller portion conditions was reported or calculatable, the range of difference was 140 kcals (portion size of a single meal manipulated) to 1865 kcals (all meals manipulated), with a median of 823 kcals.

### Risk of bias

Only a minority of studies measured daily energy intake from participant self-reports as opposed to objective researcher measured energy intake (5/14). In a limited number of studies participants were required to consume one or more meals in full (4/14) and few studies failed to address demand characteristics (4/14). Most studies reported no relevant conflicts of interest (9/14) and most studies were not pre-registered (1/14). It was rare for studies to not report on key methodological information (2/14) or fail to report or use random allocation to conditions (5/14). No studies had small sample sizes and no studies failed to use key participant eligibility criteria (e.g. currently taking appetite affecting medication). See online supplementary material table S1 for individual study risk of bias information.

### Effect of portion size condition on daily energy intake

Eighty-five effects from fourteen studies were included in the primary meta-analysis. The multi-level meta-analysis was a better fit of the data than a standard analysis (Loglikelihood ratio = 58.75, p < .001). There was a moderate-to-large reduction in daily energy intake, for smaller vs larger portions (SMD = -.709 [95% CI: -.956 to -.461], Z = 5.62, p < .001, I^2^ = 80.6%). Sensitivity analysis did not substantially influence the effect magnitude (SMDs > .624) or statistical significance of the models. Trim and fill imputed 25 effect sizes in a single level model, which did not substantially influence the effect size (SMD = -.667), and Egger’s test was significant indicative of bias (z = -14.08, p < .001), see online supplementary materials for funnel plot. When removing 13 outlying effect sizes in which the confidence intervals did not overlap with the pooled estimates (upper bound CIs < -1.03; SMDS ranged from -2.17 to -4.39), the effect size remained moderate-to-large with a small reduction in heterogeneity (SMD = -.660 [95% CI: -.860 to -.459], z = 6.43, p < .001, I^2^ = 74.9%). For meta-analysed mean difference in daily energy intake expressed as kcals, smaller portions were associated with a reduction of -235.75 [-303.02 to -168.48] calories consumed per day compared to larger portions (see Figure 2). Removal of the outlying effects did not substantially reduce this (−221.86 [95% CI: -275.69 to -.168.02]).

**Figure 2.**
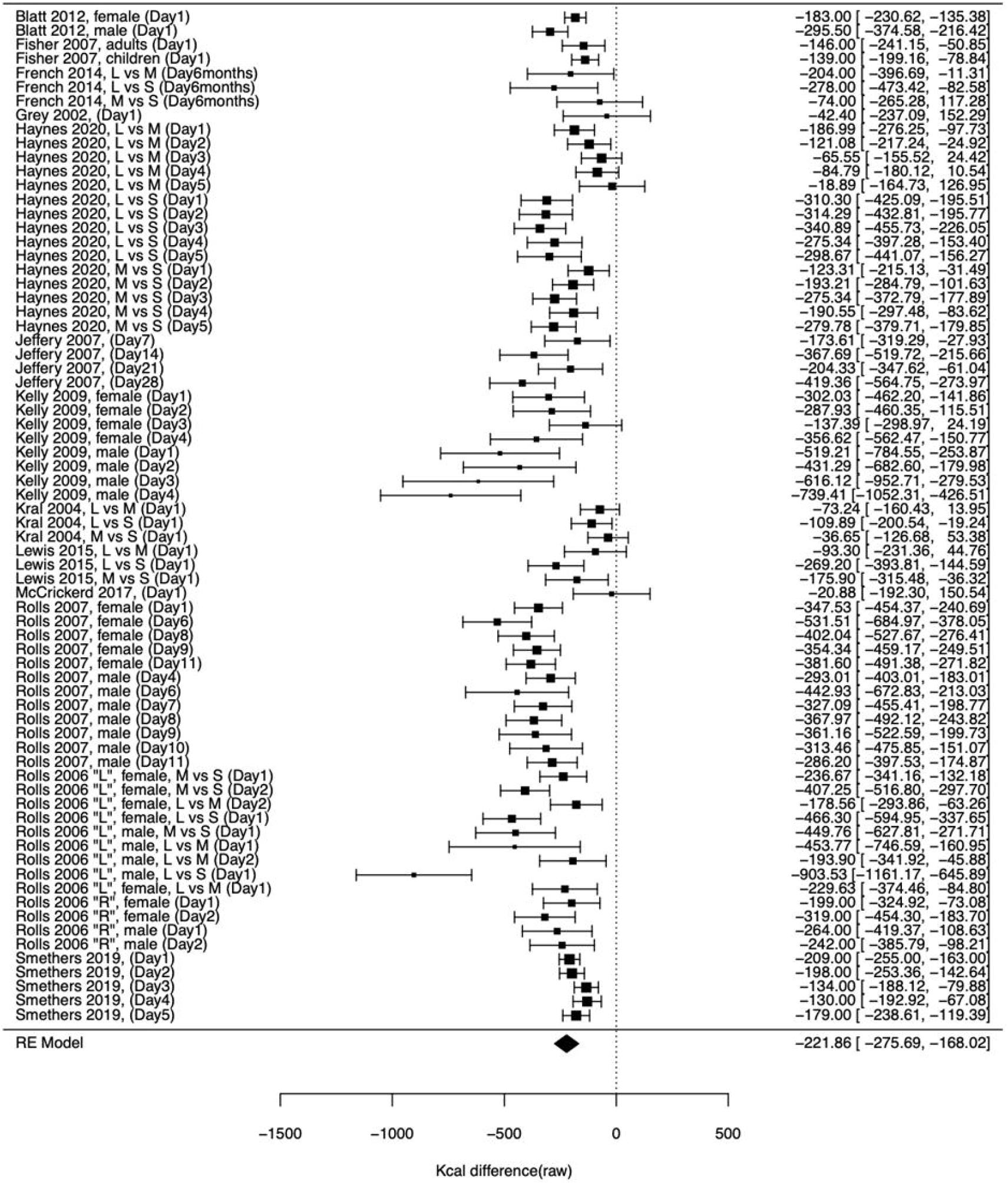
Meta-analysis of mean difference in daily energy intake between small and large portion size conditions. Figure 2 footnote: L, M and S refers to the large, medium and small portion size conditions in a study

### Participant and study features: effects on daily energy intake

#### Impact of portion size on energy intake over time

We meta-regressed the day of assessment (range day: 1 – 28, mean = 3.98, median = 2) against the effect of portion size on energy intake and there was no significant association (coefficient = -.011 [95% CI: -.038 to .016], Z = 0.81 p = .415), indicating that the influence portion size had on energy intake was not dependent on how long studies measured daily energy intake for.

#### The effect of manipulating most meals during the day vs. fewer meals

There was a significant moderation effect (X^2^(1) = 10.24, p = .001). For studies in which two or fewer meals were served as smaller vs. larger portions (30 effect sizes across 7 studies) the effect size was small-to-moderate (SMD = -.429 [95% CI: -.622 to -.228], Z = 4.23, p < .001) and the raw change in kcals was -168.23 [-233.86 to -103.61]. For studies in which more than two meals were served as smaller vs. larger portion sizes (42 effect sizes across 7 studies) there was a moderate-to-large effect size (SMD = -.900 [95% CI: -1.132 to -.669], Z = 7.63, p < .001) and the raw change in kcals was -268.53 [-335.62 to -201.44].

#### Magnitude of portion size reductions

##### Reduction of portion size (percentage)

In meta-regression, the standardised effect size was negatively associated with the magnitude portion size reduction as a percentage (coefficient = -.016 [95% CI: -.022 to -.009], Z = 4.62, p < .001), whereby based on the included studies a reduction of portion sizes served by 10% was associated with a 1.6% reduction in daily energy intake.

##### Reduction of portion size (raw kcals)

In meta-regression, the magnitude of portion size reduction expressed as a raw kcal difference between portion size conditions was negatively associated with total daily energy intake, coefficient = -0.135 ([95% CI: -0.214 to -0.056], Z = 3.56, p < .001), whereby a 100kcal total reduction in food portion size served was associated with a 14kcal reduction in daily energy intake. See Figure 3.

**Figure 3.**
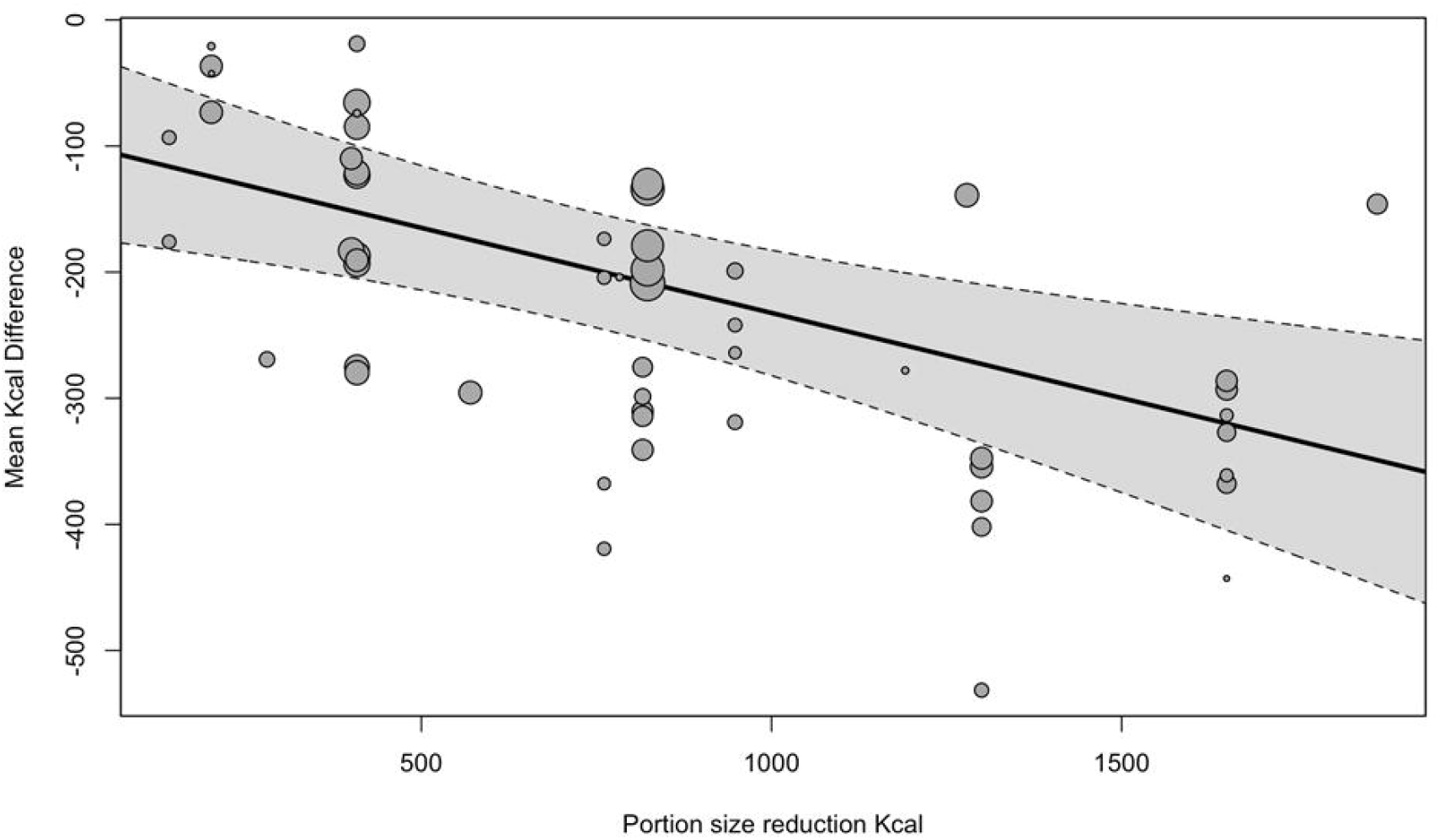
Association between difference in kcals served by portion size conditions (x axis) and daily energy intake (y axis) raw change in kcals based on portion size reduction in kcals.

### Risk of bias indicators: effects on daily energy intake

Whether energy intake was objectively measured (by the researcher) vs. self-report methods moderated the effect of portion size on daily energy intake (X^2^(1) = 4.97, p = .026). The effect of portion size on daily energy intake for studies using researcher measured energy intake (60 effect sizes from 9 studies) was SMD = -.804 ([95% CI: -1.033 to -0.575], Z = 6.87, p < .001), and for self-reported energy intake (12 effect sizes from 5 studies) was SMD = -.374 ([95% CI: -.640 to -.108], Z = 2.76, p = .00). Whether or not studies reported use of random allocation to portion size conditions did not significantly affect results (X^2^(1) = 0.02, p = .884). Whether or not a study addressed demand characteristics (X^2^(1) = 0.03, p = .867) or required participants to consume any meals in full vs. ad libitum (X^2^(1) = 0.71, p = .845) did not significantly affect results.

### Evidence for post-portion size manipulation compensatory effects

For 15 effect sizes across 7 studies that did not manipulate portion size for all meals, the impact of portion size on meal energy intake (at manipulated portion size meals) and later energy intake (at non-manipulated meals) were measured and reported separately. During the manipulated meal there was a large sized reduction for these 15 effects (SMD = -1.60 [95% CI: -2.362 to -0.841], Z = 4.13, p < .001), and the raw kcal effect on manipulated meal energy intake was -232.92 [95% CI: -357.64 to -108.21], Z = 3.66, p < .001). For non-manipulated meals following the portion size manipulated meals there was a small-to-moderate sized increase in kcals consumed after the meal in the smaller portion vs larger portion (SMD = .369 [95% CI: .024 to .714], Z = 2.10, p = .036, I^2^ = 70.5%) and the raw kcal effect was 97.72 ([95% CI 12.60 to 182.83]). Note, the standardised effect was slightly smaller in sensitivity analyses (SMD = .226 [95% CI: .010 to .442], Z = 2.05, p = .040). This analysis of a limited sub-set of studies suggests that changes to energy intake at meals caused by serving smaller vs. larger portion sizes are in part compensated by individuals eating more later in the day; approximately 42% of the reduction in energy intake observed at manipulated portion size meals was ‘compensated for’ through additional energy intake at other meals.

### Curvilinear relationship

Examining the difference in the portion size effect between large vs normal (intermediate) portions and small vs normal (intermediate) portions demonstrated a significant moderation effect (X^2^(1) = 7.57, p = .006). In large vs normal portion comparison (12 effect sizes across the 5 studies) the effect of portion size on daily energy intake was small-to-moderate in statistical size (SMD = -.389 [95% CI: -.554 to -.224], Z = 4.61, p < .001), with a raw daily kcal difference of -132.12 [95% CI: -191.92 to -72.31]. In small vs normal size portion comparisons, the effect was larger (SMD = -.578 [95% CI: -1.047 to -.109], Z = 2.43, p = .016), with raw kcal difference of -198.15 [95% CI: -331.55 to -64.75]. See Figure 4.

**Fig. 4.**
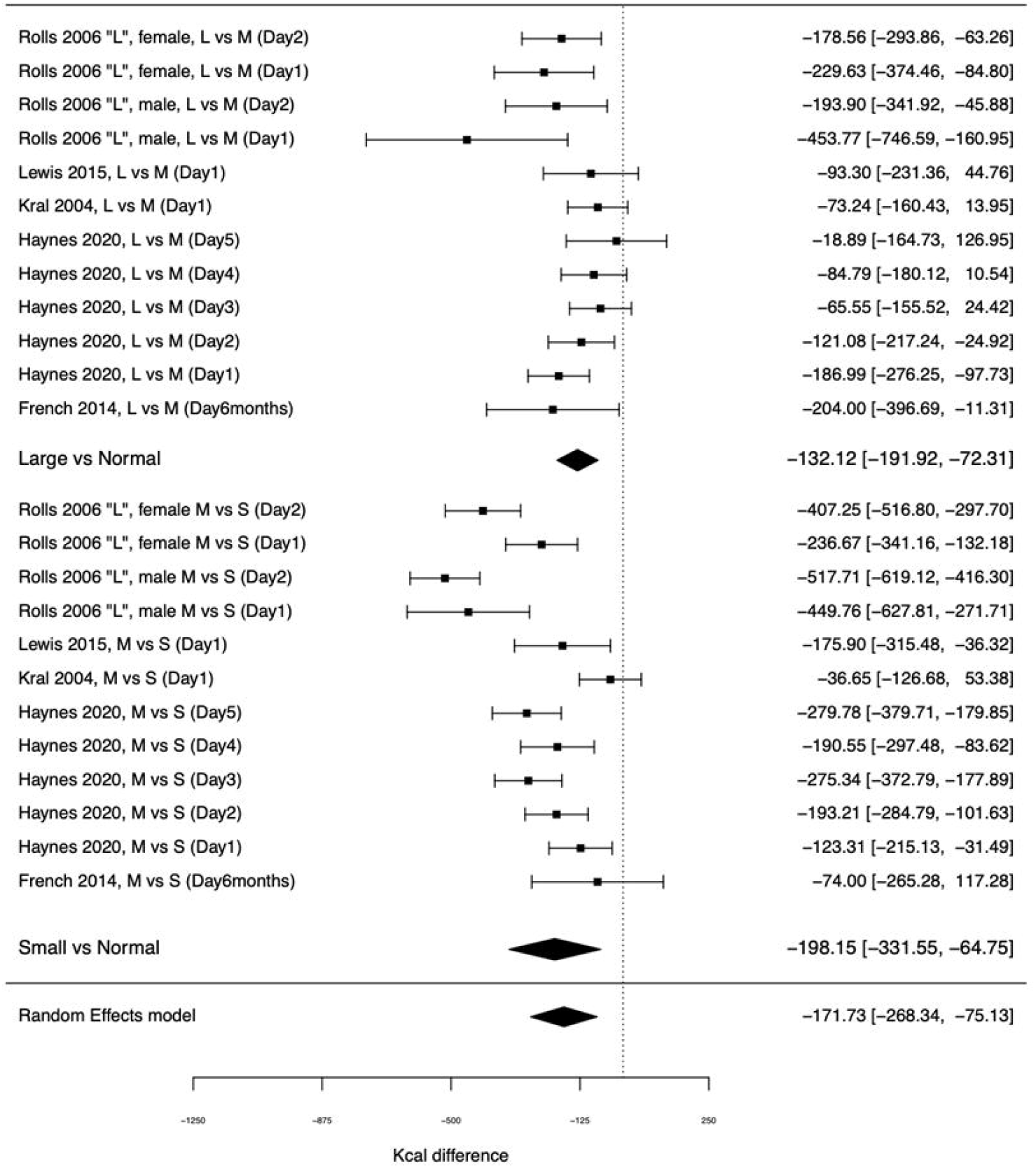
Effect of portion size on daily energy intake in studies allowing for examination of a curvilinear relationship. Figure 4 footnote: L, M and S refers to the large, medium and small portion size conditions in a study

Therefore, the impact that manipulating portion size has on daily energy intake is dependent on the size of portion that is decreased; decreasing portion size from the largest portions had a 33% smaller impact on daily energy intake than decreasing portion size of medium (intermediate) portions.

### Effect of portion size condition on body weight

Five studies examined change in body weight in smaller vs. larger portion size conditions. The standardised effect of portion size on change in body weight was SMD = .536 ([95% CI: .268 to .803], Z = 3.92, p < .001, I^2^ = 47.0%). The raw difference in change in kilograms was .579 [95% CI: .384 to .776], indicating that after allocation to being served smaller portions, participants gained 0.6 kilograms less weight than when served larger portions. See Figure 5.

**Figure 5.**
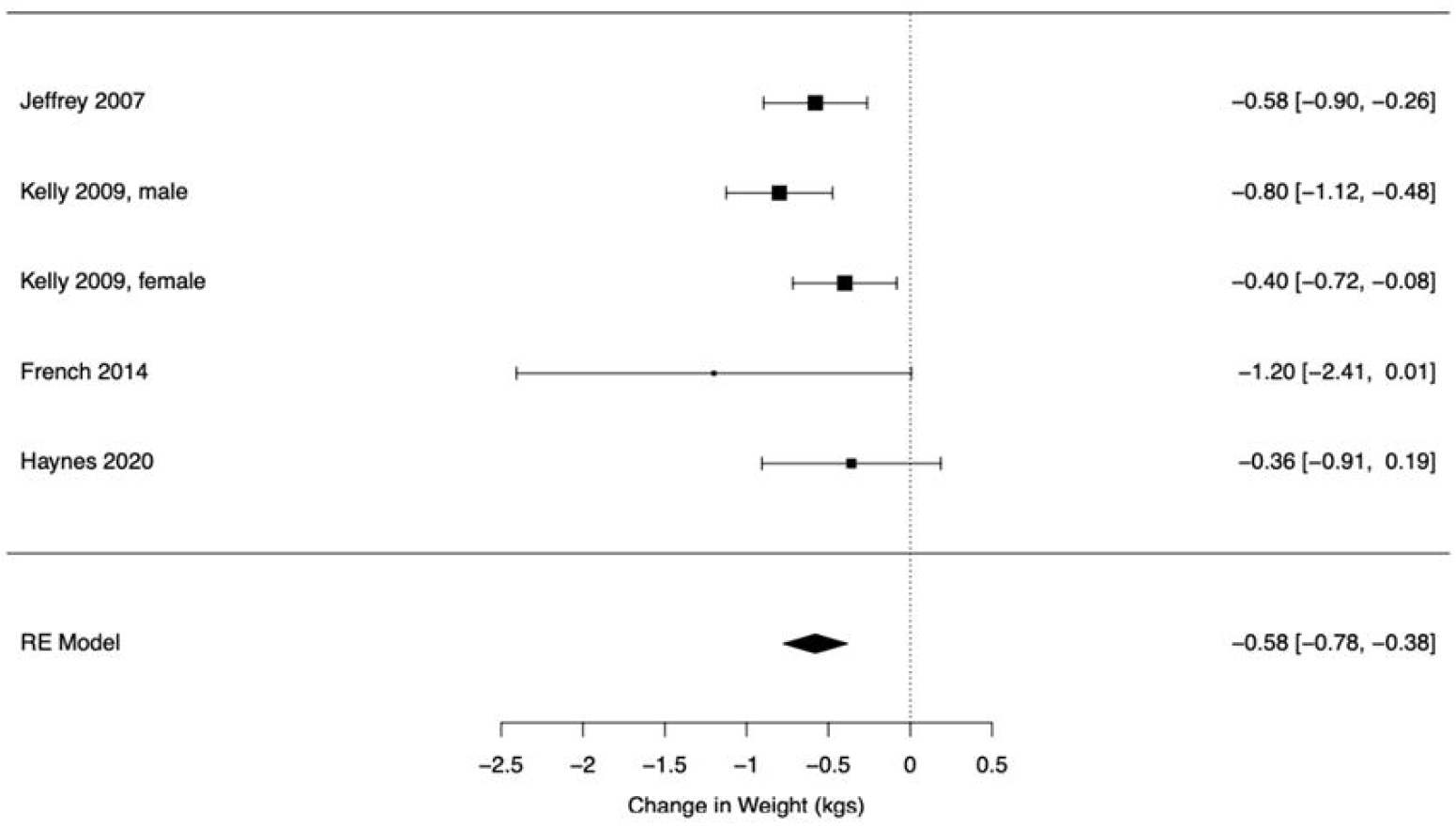
Effect of portion size condition on change in body weight.

## Discussion

We systematically reviewed and meta-analysed studies that examined the effect of experimentally manipulating food portion sizes on daily energy intake. Across fourteen eligible studies, smaller food portions resulted in lower daily energy intake and this effect was consistent across males and females. Studies varied in duration from one day to six months and there was no evidence that the effect of portion size on energy intake differed between studies that were shorter in duration or examined energy intake for longer. Reductions to daily energy intake were larger in studies that manipulated the portion size of foods at most meals as opposed to studies that only manipulated portion size at one or two meals. Larger reductions to served food portion sizes (expressed as either a % decrease in portion size from the largest portion or difference in kcals served) resulted in larger changes to daily energy intake. There was some evidence of compensation (e.g. participants eating more later in the day) when served smaller vs. larger portions, but this compensation was only partial. Studies tended to be relatively low in risk of bias and there was minimal evidence that studies higher in risk of bias (e.g. did not report use of random allocation to portion size conditions) produced different results to studies not exhibiting risk of bias. However, studies that relied in part on participant self-reports of food consumed to calculate energy intake reported smaller effects of portion size on daily energy intake than studies relying on researcher measured energy intake. Given that participant self-reported energy intake is prone to recall bias and inaccuracy (33), participant reporting biases may underestimate the effect of portion size on energy intake in some studies.

Meta-analyses of the effect of portion size on energy intake have been limited predominantly to studies measuring energy intake at a single acute meal, to date. In a meta-analysis of studies sampling children, larger (vs. smaller) portion sizes were estimated to have a moderate-sized statistical effect on energy intake (SMD = 0.47) (7). In a meta-analysis consisting of adults and children (10), increasing portion size by 100% resulted in on average a 35% increase in meal energy intake (or in reverse a 50% portion size reduction being associated with a 19% decrease in meal energy intake). After accounting for potential publication bias, the effect of decreasing portion size on daily energy intake in the present meta-analysis was a statistically moderate sized effect and based on the results of analyses of all included studies, a 50% reduction in portion sizes would be associated with an 8% decrease in daily energy intake, or expressed as energy; a 100kcal total reduction in portion sizes served would be estimated to result in approximately 14kcal decrease to daily energy intake. However, consistent with short-term studies of the effect of portion size on acute energy intake (10, 32), we found evidence from a small sub-set of studies (N=5) that the effect of portion size on daily energy intake was curvilinear; reductions to daily energy intake were markedly smaller (∼33%) when reducing portion size from a large portions to a ‘normal’/intermediate portion, compared to reducing portion size from a ‘normal’/ intermediate portion to a smaller portion. In other words, the extent to which daily energy intake appears to ‘compensate’ for energy served is far greater when portion size is very large. This is important because most studies included in the present review served participants very large amounts of food in the ‘large’ portion size condition and these portions are unlikely to be representative of portion sizes served in everyday life. For example, results from a laboratory study (19) examining the effect of very large portions (i.e. serving participants in excess of 6000kcals per day) found that a 2050kcal difference in energy served per day between the larger and smaller portion conditions of the study resulted only in a 419 difference in average daily energy intake (80% ‘compensation’ in energy consumed compared to energy served). Conversely in a different laboratory study that compared meals that were chosen to be perceived as being ‘normal’ in size (i.e. typical of everyday portion sizes) vs. smaller portioned meals (18), a 408kcal difference in energy served across the day resulted in far less ‘compensation’; a 210kcal decrease in average daily energy intake (only 49% compensation). Therefore, caution should be taken and curvilinear relations accounted for when extrapolating the results of our main analyses to estimate how much reducing portion sizes in everyday life would be expected to decrease daily energy intake.

The present results provide convincing evidence that food portion size has a causal impact on daily energy intake. We assume that portion size impacts on daily energy intake because there appears to be a lack of tight short-term control of energy intake in humans (34) and food intake behaviour is context dependent, whereby individuals can easily eat more or less food dependent on the absence vs. presence of environmental cues or factors, such as portion size. Consistent with other studies (16), we found evidence that there is some energy intake compensation in response to manipulations of portion size (e.g. eating more/less after having been served a smaller/larger portion size), but this compensation was only partial (approximately 42%). Furthermore, analyses suggested that this compensation does not become larger over time (i.e. increased compensation after several days of being served small portions as opposed to one day). The compensation in response to smaller vs. larger portions that occurs each day but does not become larger over time may be explained by the short-term physiological regulation of food intake being largely determined by emptiness of the gut (34) (i.e. why smaller portions may promote some short-term increase in energy intake on the same day) and that cognitive regulation of food intake is episodic memory specific and therefore influenced only by recent eating episodes (35, 36) (i.e. during the same day).

Because portion sizes of some commercially provided foods are known to have increased in recent times (2), the present findings suggest that this is likely to have contributed to increases in population level energy intake and the prevalence of overweight and obesity. There have been some questions raised over the lack of causal evidence on the effect of portion size on body weight and therefore the public health benefit of reducing portion sizes (5). We meta-analysed a small number of studies that measured participant body weight and found that larger portions were associated with greater weight gain than smaller portions. Collectively the present findings indicate that reductions to food portion sizes may reduce population level prevalence of overweight and obesity.

We were limited to examining only gender as a moderating participant characteristic of the effect of portion size on daily energy intake and found no evidence of moderation. However, this sub-group analysis consisted of a small number of effects and therefore should be interpreted with caution. We were unable to examine whether participant BMI moderated the effect of portion size on daily energy intake, or the potential moderating effect of individual differences in trait eating behaviours, such as satiety responsiveness (37). Studies to date have not found convincing evidence for participant characteristics that consistently moderate the effect of portion size on energy intake (38). However, it will be important for future research to address this and examine if the impact that reducing portion size has on daily energy intake is beneficial to the majority of the population or if there are individual level factors determine responsiveness to portion size. A further limitation is due to the studies available we were unable to examine whether properties or presentation of food determine the effect of portion size on daily energy intake. There was suggestive evidence of publication bias and some of the included studies scored high for markers of risk of bias. Analyses accounting for publication bias still resulted in a significant (but slightly smaller) effect of portion size on energy intake and results were consistent in analyses that accounted for risk of bias. Effect sizes were largely from adult studies and therefore may not be generalisable to children, although similar findings have been demonstrated in children. As noted, the number of eligible studies was relatively small and therefore caution should be taken in the interpretation of some of the reported sub-group analyses. Most studies were short in duration and measured energy intake for 1-2 days, therefore further studies examining the effect portion size on daily energy intake and body weight over longer time periods would be valuable. A further limitation is that the majority of studies were laboratory based and therefore may not be reflective of real-world eating due to social desirability concerns (39, 40). A recent study found that the effect of portion size on short-term energy intake was larger when tested in the real-world vs. laboratory (41), therefore we presume that the reliance on laboratory based studies in the present meta-analysis would be more likely to under rather than overestimate the effect of portion size on daily energy intake.

### Conclusions

Smaller food portion sizes substantially decrease daily energy intake and there is evidence that over time this results in lower body weight. Reducing food portion sizes may be an effective population level strategy to reduce obesity.

## Data Availability

Data described in the manuscript, code book, and analytic code is publicly and freely available without restriction at https://osf.io/dj4yf/

## Author Contributions

ER designed the research, conducted the research, had primary responsible for the final content and wrote the paper. AJ designed the research, conducted the research, analysed data and wrote the paper. IML and ZP conducted the research.

## References

1. Livingstone MBE, Pourshahidi LK. Portion size and obesity. Advances in nutrition. 2014;5(6):829–34.

2. Young LR, Nestle M. The contribution of expanding portion sizes to the US obesity epidemic. American journal of public health. 2002;92(2):246–9.

3. Ledikwe JH, Ello-Martin JA, Rolls BJ. Portion sizes and the obesity epidemic. The Journal of nutrition. 2005;135(4):905–9.

4. Robinson E, Jones A, Whitelock V, Mead BR, Haynes A. (Over) eating out at major UK restaurant chains: observational study of energy content of main meals. bmj. 2018;363.

5. Benton D. Portion size: what we know and what we need to know. Crit Rev Food Sci Nutr. 2015;55(7):988–1004.

6. Muc M, Jones A, Roberts C, Sheen F, Haynes A, Robinson E. A bit or a lot on the side? Observational study of the energy content of starters, sides and desserts in major UK restaurant chains. BMJ open. 2019;9(10):e029679.

7. Reale S, Hamilton J, Akparibo R, Hetherington M, Cecil J, Caton S. The effect of food type on the portion size effect in children aged 2–12 years: A systematic review and meta-analysis. Appetite. 2019;137:47–61.

8. Sheen F, Hardman CA, Robinson E. Plate-clearing tendencies and portion size are independently associated with main meal food intake in women: A laboratory study. Appetite. 2018;127:223–9.

9. Rolls BJ, Roe LS, Meengs JS, Wall DE. Increasing the portion size of a sandwich increases energy intake. Journal of the American Dietetic Association. 2004;104(3):367–72.

10. Zlatevska N, Dubelaar C, Holden SS. Sizing up the effect of portion size on consumption: a meta-analytic review. Journal of Marketing. 2014;78(3):140–54.

11. Marteau TM, Hollands GJ, Shemilt I, Jebb SA. Downsizing: policy options to reduce portion sizes to help tackle obesity. BMJ. 2015;351.

12. Robinson E, Henderson J, Keenan GS, Kersbergen I. When a portion becomes a norm: Exposure to a smaller vs. larger portion of food affects later food intake. Food quality and preference. 2019;75:113–7.

13. Robinson E, Kersbergen I. Portion size and later food intake: evidence on the “normalizing” effect of reducing food portion sizes. The American journal of clinical nutrition. 2018;107(4):640–6.

14. Raghoebar S, Haynes A, Robinson E, Van Kleef E, De Vet E. Served portion sizes affect later food intake through social consumption norms. Nutrients. 2019;11(12):2845.

15. Haynes A, Hardman CA, Makin AD, Halford JC, Jebb SA, Robinson E. Visual perceptions of portion size normality and intended food consumption: A norm range model. Food quality and preference. 2019;72:77.

16. Haynes A, Hardman CA, Halford JC, Jebb SA, Robinson E. Portion size normality and additional within-meal food intake: two crossover laboratory experiments. British Journal of Nutrition. 2020;123(4):462–71.

17. Lewis HB, Ahern AL, Solis[Trapala I, Walker CG, Reimann F, Gribble FM, et al. Effect of reducing portion size at a compulsory meal on later energy intake, gut hormones, and appetite in overweight adults. Obesity. 2015;23(7):1362–70.

18. Haynes A, Hardman CA, Halford JC, Jebb SA, Mead BR, Robinson E. Reductions to main meal portion sizes reduce daily energy intake regardless of perceived normality of portion size: a 5 day cross-over laboratory experiment. International Journal of Behavioral Nutrition and Physical Activity. 2020;17(1):21.

19. Rolls BJ, Roe LS, Meengs JS. The effect of large portion sizes on energy intake is sustained for 11 days. Obesity. 2007;15(6):1535–43.

20. French SA, Mitchell NR, Wolfson J, Harnack LJ, Jeffery RW, Gerlach AF, et al. Portion size effects on weight gain in a free living setting. Obesity. 2014;22(6):1400–5.

21. Kelly MT, Wallace JM, Robson PJ, Rennie KL, Welch RW, Hannon-Fletcher MP, et al. Increased portion size leads to a sustained increase in energy intake over 4 d in normal-weight and overweight men and women. British journal of nutrition. 2009;102(3):470–7.

22. Jeffery RW, Rydell S, Dunn CL, Harnack LJ, Levine AS, Pentel PR, et al. Effects of portion size on chronic energy intake. International Journal of Behavioral Nutrition and Physical Activity. 2007;4(1):27.

23. Blundell J, De Graaf C, Hulshof T, Jebb S, Livingstone B, Lluch A, et al. Appetite control: methodological aspects of the evaluation of foods. Obesity Reviews. 2010;11(3):251–70.

24. Higgins JP, Altman DG, Gøtzsche PC, Jüni P, Moher D, Oxman AD, et al. The Cochrane Collaboration’s tool for assessing risk of bias in randomised trials. Bmj. 2011;343.

25. Robinson E, Bevelander K, Field M, Jones A. Methodological and reporting quality in laboratory studies of human eating behavior. Appetite. 2018;125:486–91.

26. Kersbergen I, Whitelock V, Haynes A, Schroor M, Robinson E. Hypothesis awareness as a demand characteristic in laboratory-based eating behaviour research: An experimental study. Appetite. 2019;141:104318.

27. Pastor DA, Lazowski RA. On the Multilevel Nature of Meta-Analysis: A Tutorial, Comparison of Software Programs, and Discussion of Analytic Choices. Multivariate Behavioral Research. 2018;53(1):74–89.

28. Viechtbauer W, Cheung MWL. Outlier and influence diagnostics for meta[analysis. Research synthesis methods. 2010;1(2):112–25.

29. Egger M, Smith GD, Schneider M, Minder C. Bias in meta-analysis detected by a simple, graphical test. BMJ. 1997;315(7109):629–34.

30. Duval S, Tweedie R. Trim and fill: a simple funnel plot–based method of testing and adjusting for publication bias in meta[analysis. Biometrics. 2000;56(2):455–63.

31. Alcohol Health Alliance UK (2020). Drinking in the dark: How alcohol labelling fails consumers. https://ahauk.org/wp-content/uploads/2020/08/DRINKING-IN-THE-DARK.pdf.

32. Roe LS, Kling SMR, Rolls BJ. What is eaten when all of the foods at a meal are served in large portions? Appetite. 2016;99:1–9.

33. Ravelli MN, Schoeller DA. Traditional Self-Reported Dietary Instruments Are Prone to Inaccuracies and New Approaches Are Needed. Frontiers in Nutrition. 2020;7(90).

34. Rogers PJ, Brunstrom JM. Appetite and energy balancing. Physiol Behav. 2016;164(Pt B):465–71.

35. Higgs S, Robinson E, Lee M. Learning and memory processes and their role in eating: implications for limiting food intake in overeaters. Current obesity reports. 2012;1(2):91–8.

36. Robinson E, Aveyard P, Daley A, Jolly K, Lewis A, Lycett D, et al. Eating attentively: a systematic review and meta-analysis of the effect of food intake memory and awareness on eating. The American journal of clinical nutrition. 2013;97(4):728–42.

37. Zuraikat FM, Smethers AD, Rolls BJ. Potential moderators of the portion size effect. Physiology & Behavior. 2019;204:191–8.

38. Robinson E, Haynes A. Individual differences and moderating participant characteristics in the effect of reducing portion size on meal energy intake: Pooled analysis of three randomized controlled trials. Appetite. 2021;159:105047.

39. Robinson E, Kersbergen I, Brunstrom JM, Field M. I’m watching you. Awareness that food consumption is being monitored is a demand characteristic in eating-behaviour experiments. Appetite. 2014;83:19–25.

40. 1. Robinson E, Proctor M, Oldham M, Masic U. The effect of heightened awareness of observation on consumption of a multi-item laboratory test meal in females. Physiology & behavior. 2016;163:129–35.

41. Gough T, Haynes A, Clarke K, Hansell A, Kaimkhani M, Price B, et al. Out of the lab and into the wild: The influence of portion size on food intake in laboratory vs. real-world settings. Appetite. 2021;162:105160.

42. Blatt AD, Williams RA, Roe LS, Rolls BJ. Effects of Energy Content and Energy Density of Pre-Portioned Entrées on Energy Intake. Obesity. 2012;20(10):2010–8.

43. Fisher JO, Arreola A, Birch LL, Rolls BJ. Portion size effects on daily energy intake in low-income Hispanic and African American children and their mothers. Am J Clin Nutr. 2007;86(6):1709–16.

44. Gray RW, French SJ, Robinson TM, Yeomans MR. Dissociation of the effects of preload volume and energy content on subjective appetite and food intake. Physiology & Behavior. 2002;76(1):57–64.

45. Kral TV, Roe L, Rolls BJ. Combined effects of energy density and portion size on energy intake in women. Am J Clin Nutr. 2004.

46. McCrickerd K, Lim CM, Leong C, Chia-Ming E, Forde C. Texture-Based Differences in Eating Rate Reduce the Impact of Increased Energy Density and Large Portions on Meal Size in Adults. journal of nutrition Ingestive Behavior and Neurosciences. 2017.

47. Rolls BJ, Roe LS, Meengs JS. Larger portion sizes lead to a sustained increase in energy intake over 2 days. J Am Diet Assoc. 2006;106(4):543–9.

48. Rolls BJ, Roe LS, Meengs JS. Reductions in portion size and energy density of foods are additive and lead to sustained decreases in energy intake. Am J Clin Nutr. 2006;83(1):11–7.

49. Smethers AD, Roe LS, Sanchez CE, Zuraikat FM, Keller KL, Kling SMR, et al. Portion size has sustained effects over 5 days in preschool children: a randomized trial. Am J Clin Nutr. 2019;109(5):1361-72.1. 25

