## Supplementary material for "Downsizing food: A systematic review and meta-analysis examining the effect of reducing served food portion sizes on daily energy intake and body weight": Online supplementary material

Online Supplementary Materials

Example of search terms used

PubMed: (Energy density OR portion size OR food reformulation) AND (energy intake OR calories OR food intake OR appetite OR eating). Filter: Humans. Records identified = 5,880

Deviations from planned analyses

We had originally planned to systematically review and meta-analyse studies that had manipulated portion size of foods and studies that had manipulated energy density of foods in the same project. However, after conducting searches and identifying eligible articles we determined that the scope of the review is too large to be completed in line with the original project timeline, as a result of staffing during the COVID pandemic and a larger number of eligible energy density studies than anticipated. For these reasons, we do not include energy density studies in this project and will return to energy density studies in a separate piece of research at a later date. This decision was made during piloting of data extraction. The only changes to the pre-registered protocol is the exclusion of studies that have manipulated energy density of food only (and not portion size) and therefore study type (i.e. portion size vs. energy density) was not examined as a sub-group factor in planned quantitative analyses.

Planned influential case and publication bias analyses

We characterised outliers as any effect sizes for which the upper bound of the 95% confidence interval was lower than the lower bound of the pooled effect confidence interval (i.e., extremely small effects) or for which the lower bound of the 95% confidence interval was higher than the upper bound of the pooled effect confidence interval (i.e., extremely large effects). If any outliers were identified then we planned to report the results of meta-analyses with the outliers removed. To address influential cases, we computed DFBETAS values for each effect size. Influential cases were identified if DFBETAS values > 1 (indicative of a >1 change in the standard deviation of the estimated co-efficient after removal of the study) (1). To increase sensitivity, we conducted leave-one-out analyses by removing each study (k) from the analyses, and refitting the model. If the removal of k substantially influences the model (statistical significance of the model changes from p < .05 to p > .05 (or p >.05 to p < .05), this was classed as an influential case. We examined evidence for publication bias in the primary analyses by examining asymmetry of effect sizes. We plotted and visually inspected funnel plots. Next, we conducted an Egger’s test of asymmetry and Trim and Fill procedure. For Egger’s test (2), if the intercept is significantly different from 0 at p >.10 this is indicative of bias. Trim and Fill (3) removes less precise studies which might case any asymmetry (‘trim’), re-estimates the overall effect size, and then replaces removed studies and missing counterparts (‘fill’) required for symmetry. We planned to report i) the number of missing (‘filled’) studies, and ii) the estimate of the effect size following their inclusion.

*Online Supplementary Figure 1: Funnel plot of the effect sizes from the primary analyses with outliers removed*

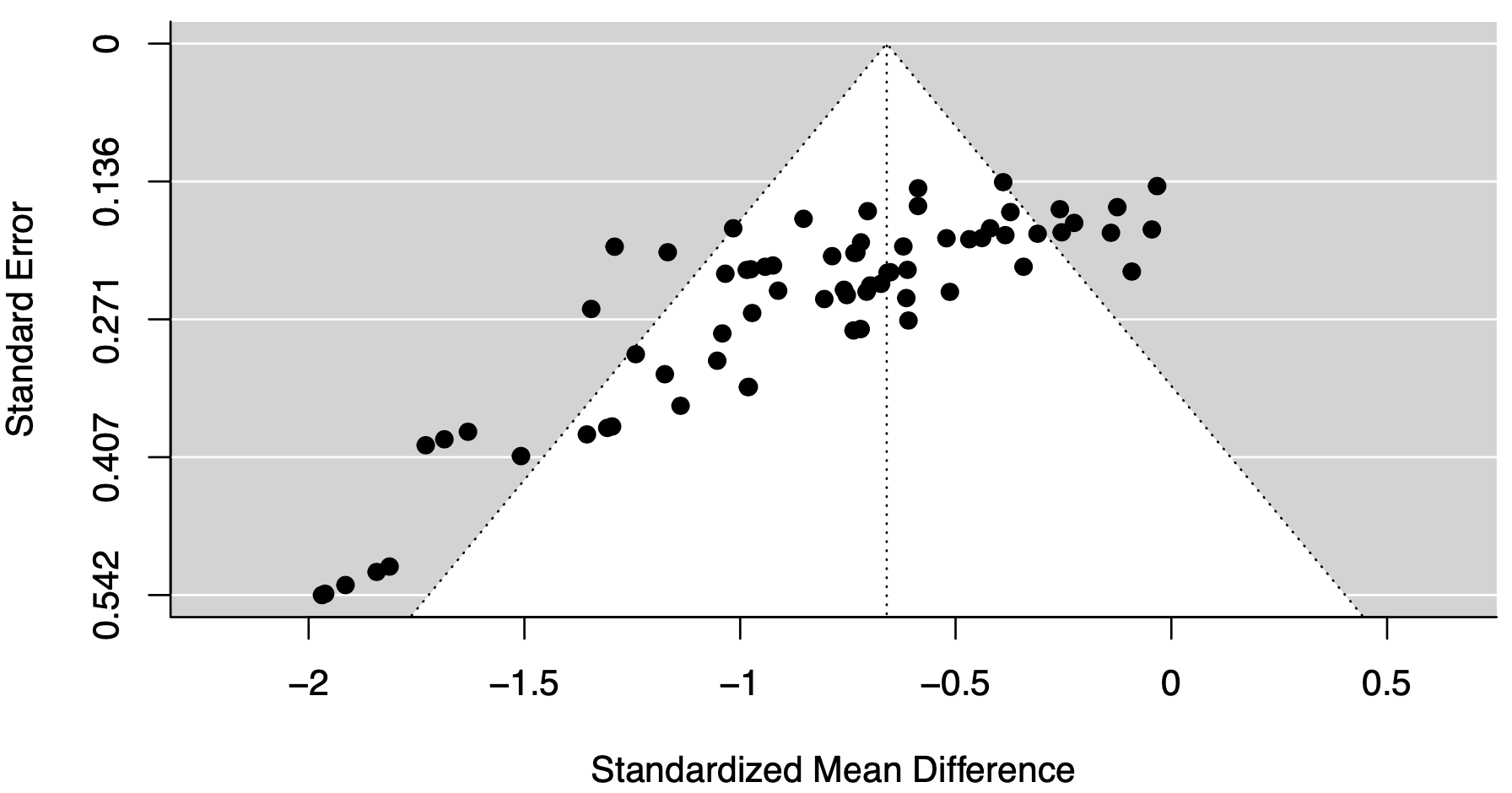

References

1.	Viechtbauer W, Cheung MWL. Outlier and influence diagnostics for meta‐analysis. Research synthesis methods 2010;1(2):112-25.

2.	Egger M, Smith GD, Schneider M, Minder C. Bias in meta-analysis detected by a simple, graphical test. Bmj 1997;315(7109):629-34.

3.	Duval S, Tweedie R. Trim and fill: a simple funnel‐plot–based method of testing and adjusting for publication bias in meta‐analysis. Biometrics 2000;56(2):455-63.

*Table S1. Risk of bias information for included studies*

| **Study** | **Self-reported energy intake used** | **Key exclusion criteria omitted or unclear** | **Lack of key method information** | **Unclear or lack of random allocation to conditions** | **Participants required to consume meal(s) in full** | **Demand characteristics not addressed or unclear** | **Small sample size** | **Study was not pre-registered** | **Reported relevant conflicts of interest or information missing** |
| --- | --- | --- | --- | --- | --- | --- | --- | --- | --- |
| Blatt 2012 | N | N | N | Y | Y | N | N | Y | N |
| Fisher, 2007 | N | N | N | N | N | Y | N | Y | Y |
| French, 2014 | Y | N | N | N | N | Y | N | Y | N |
| Gray, 2002 | Y | N | N | Y | Y | N | N | Y | Y |
| Haynes, 2020 | N | N | N | N | N | N | N | N | Y |
| Jeffery, 2007 | Y | N | N | N | N | Y | N | Y | N |
| Kelly, 2009 | N | N | Y | N | N | N | N | Y | N |
| Kral, 2004 | N | N | N | Y | Y | N | N | Y | N |
| Lewis, 2015 | Y | N | N | N | Y | N | N | Y | N |
| McCrickerd, 2017 | Y | N | N | N | N | N | N | Y | N |
| Rolls 2006 ‘Larger’ | N | N | Y | Y | N | N | N | Y | Y |
| Rolls 2006 ‘Reductions’ | N | N | N | Y | N | N | N | Y | N |
| Rolls, 2007 | N | N | N | N | N | N | N | Y | Y |
| Smethers, 2019 | N | N | N | N | N | Y | N | Y | N |
